# Distinct lipid mediator profiles in brain and liver tissues in Alzheimer’s disease

**DOI:** 10.64898/2026.09.22.26363708

**Authors:** Sitong Zhou, Kamil Borkowski, Nuanyi Liang, Thomas G. Beach, Geidy E. Serrano, Kyoungmi Kim, John W. Newman, Izumi Maezawa, Lee-Way Jin

## Abstract

**INTRODUCTION:** Inflammation plays a central role in Alzheimer’s Disease (AD), with oxylipins, endocannabinoids, and polyunsaturated fatty acids (PUFAs) acting as potent modulators. Soluble epoxide hydrolase (sEH), an enzyme with proinflammatory products, is upregulated in the AD brain. Emerging evidence suggests that AD-related inflammation may reflect systemic metabolic changes, however, peripheral organs remain understudied. Because the liver has a pivotal role in lipid metabolism and bioactive lipid mediator homeostasis, we hypothesized that inflammatory lipid mediator changes in the AD brain may be reflected in, and potentially shaped by, hepatic lipid metabolism.

**METHODS:** Paired postmortem brain and liver tissues from 16 cognitively normal and 63 AD donors with median postmortem intervals (PMI) of 3.08 hours were analyzed for free and esterified forms of oxylipins and endocannabinoids. ANCOVA models evaluated effects of AD diagnosis, sex, and AD-by-sex interactions, adjusting for analytical batch, age, body mass index, and PMI.

**RESULTS:** AD-associated differences in lipid mediator profiles were observed in both brain and liver, with a broader pattern of differences in liver. Differences involved CYP/sEH-related metabolites and fatty acid ethanolamides, with several metabolites also showing AD-by-sex interactions. Sex-associated differences were also more pronounced in the liver than the brain.

**DISCUSSION:** These results suggest AD-associated lipid dysregulation across both central and peripheral tissues. The prominent hepatic alterations highlight the liver as an important site of peripheral lipid mediator dysregulation in AD and suggest that hepatic metabolic changes may contribute to or reflect inflammatory lipid remodeling in the brain.

## 1. Introduction

Inflammation is a key component of neurodegenerative diseases like Alzheimer’s disease (AD) and often reflected in altered levels of biologically active oxygenated products of polyunsaturated fatty acids (PUFAs), i.e., oxylipins, endocannabinoids and endocannabinoid-like substances (Oxyl-EC). The structures of representative PUFA-derived oxylipins are shown in **Figure 1**. Within this metabolic system, soluble epoxide hydrolase (sEH) is crucial due to its activities in the hydrolysis of pro-resolving epoxy-fatty acids (epoxides) to pro-inflammatory dihydroxy-fatty acids (diols) [1–4]. sEH is an enzyme highly expressed in both periphery tissues and the central nervous system (CNS). Upregulation of sEH in the brain has been reported in various CNS disease models including AD [1, 5–7]. Moreover, sEH pathway and fatty acids ethanolamides (FAEA) in plasma and cerebrospinal fluid (CSF) were associated with AD pathology, early cognitive decline, cognitive resilience to Tau and amyloid beta (Aβ) pathology and several processes that contribute to AD development [8–11]. Additionally, midchain alcohols from lipoxygenases and autoxidative processes are also associated with pro-inflammatory activities [12]. Collectively, the involvement of Oxyl-EC in AD pathology is established, yet extensive investigation is necessary to precisely understand the inflammatory process [13–16].

**Figure 1.**
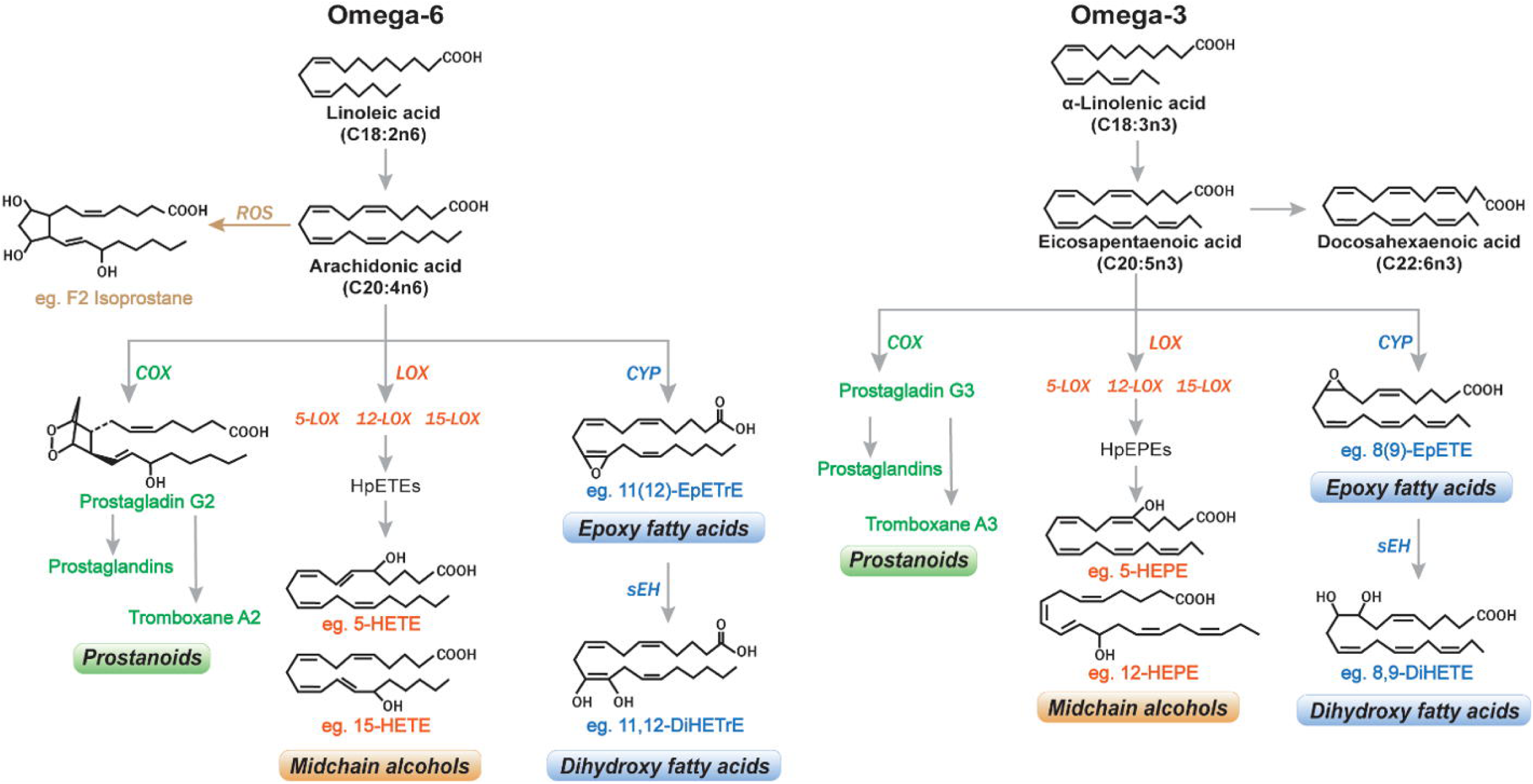
Representative oxylipins derived from linoleic acid (LA) or α-linolenic acid (αLA). Unsaturated fatty acid (e.g., LA or αLA) can be transformed into a variety of structures including prostanoids, midchain alcohols, epoxy fatty acids, and dihydroxy fatty acids, etc. The interactions involve with cytochrome 450s (CYPs), epoxide hydrolases (EH), lipoxygenases (LOX), cyclooxygenases (COX), reactive oxygen species (ROS) and other enzymes. These reactions result in the formation of epoxides (e.g., epoxyeicosatrienoic acids (EpETrEs)), diols (e.g., dihydroxyeicosatrienoic acids (DiHETrEs)), mono-alcohols (e.g., hydroxyeicosatrienoic acids (HETEs)), and isomeric prostaglandins (e.g., F2 isoprostanes), etc.

To date, AD-related oxylipin differences have been observed in mouse models, human plasma, CSF, and postmortem brain tissue [6, 7, 14, 17, 18]. However, most studies only target the free (unesterified) oxylipins, while the majority of oxylipins (∼90%) are esterified (bound) in the brain and plasma [19–21]. Generally, free oxylipins are associated with proteins or particles, whereas bound oxylipins are esterified into complex lipids, such as cellular phospholipids, cholesterol esters, and lipoproteins. These lipoproteins serve as the primary plasma transporters of oxylipins [19, 22]. Since the esterified oxylipin pool serves as a reservoir and modulator for free oxylipins to be released or re-esterified, it is essential to investigate the roles of both forms of oxylipins in AD pathology [23, 24].

While AD research has conventionally centered on the brain, these inflammatory processes are not confined to central compartment but extend to peripheral systems including the liver [25–28]. The essential crosstalk between the periphery and brain is increasingly recognized as reflecting a coordinated systemic metabolic regulation rather than a purely brain-centered processes [29–31]. The blood-brain barrier (BBB) is a physiological interface involved in the regulatory exchange of inflammatory mediators and immune cells [32, 33]. BBB leakage is implicated in AD, and its dysfunction permits peripheral inflammation-related cytokines to infiltrate the brain and further contribute to neuroinflammation [34–36]. Notably, peripheral inflammatory cytokine levels correlate with AD progression [36, 37]. Moreover, while Aβ is considered the hallmark of AD, the liver is involved in blood Aβ clearance and brain Aβ regulation. [38, 39]. Hepatic sEH activity has also been implicated in the regulation of systemic Aβ metabolism [40]. These findings indicate that AD is not solely a brain disorder, but rather a systemic metabolic disease involving coordinated lipid inflammatory dysregulation across central and peripheral organs. However, the specific pathological changes associated with peripheral abnormalities remain unclear, and the AD pathology from the hepatic perspective is yet to be established.

To address the knowledge gap, we aimed to investigate the involvement of peripheral and central lipid metabolism in AD pathogenesis. We quantified Oxyl-EC from paired postmortem brain and liver tissues in comparison of cognitively normal and AD cases. Notably, to our knowledge, this is the first in-depth analysis of both esterified and non-esterified (free) oxylipin pools in the effort to obtain a comprehensive characterization of lipid mediator metabolism across paired central and peripheral tissues. By comparing these metabolite profiles, we sought to understand whether hepatic lipid metabolic alterations are associated with inflammatory lipid remodeling in the AD brain. We hypothesized that the liver is not merely affected by AD pathology but represents an active component of a brain-liver lipid metabolic axis that contributes to central inflammatory processes.

## 2. Materials and Methods

### 2.1 Human tissue samples

Frozen prefrontal cortex and liver samples were obtained from the Banner Sun Health Research Institute (BSHRI) Brain and Body Donation Program (BBDP, www.brainandbodydonationprogram.org), where written informed consent was obtained and approved by the BSHRI Review Board, and participants were evaluated through standardized procedures as previously reported [41]. The study was conducted in accordance with the Declaration of Helsinki and approved by the Institutional Review Board of WCG (Study Number 1132516 08/05/2022).

### 2.2 Subject selection

The study group consisted of 79 Caucasian subjects with a 3.08-hour median postmortem interval (PMI) for tissue collection. Among the subjects, 16 were cognitively normal (control) (male: N = 9; female: N = 7), and 63 were diagnosed with AD (male: N = 28; female: N = 35). Sample characteristics such as age and body mass index (BMI) were summarized in **Table 1**. We obtained paired brain and liver tissues from the same subjects; however, in six cases, the brain tissue was unavailable. In the effort to reduce patient heterogeneity in the study cohort, only individuals with homozygous allele apolipoprotein E (ApoE3/3 and ApoE4/4) were included, while ApoE3/4 individuals were excluded to avoid potential heterogeneity associated with mixed ApoE alleles.

**Table 1.** Characteristics of cognitively normal and AD subjects. (BMI, body mass index.)

|  | **Cognitively normal (N = 16)** | **AD (N = 63)** |
| --- | --- | --- |
| **Age (years)** |  |  |
| Mean (SD) | 81.5 (13.7) | 85.9 (7.7) |
| Range (min – max) | 53 - 99 | 59 - 101 |
| **Sex** |  |  |
| Male | 9 (56%) | 28 (44%) |
| Female | 7 (44%) | 35 (56%) |
| **BMI (kg/m<sup>2</sup>)** |  |  |
| Mean (SD) | 25.2 (0.51) | 24.4 (0.37) |
| Range (min – max) | 22.2 – 32.5 | 14.5 – 36.7 |

### 2.3 Extraction of esterified oxylipin, endocannabinoids, and PUFAs

Frozen tissue samples (20 mg ∼ 50 mg) were sectioned and weighed on dry ice. Similar to previously reported, tissue samples were spiked with 5 µL of 0.2 mg/mL butylated hydroxyl toluene (BHT) ethylene diamine tetraacetic acid (EDTA) in 1:1 methanol: water. After the addition of 4mm stainless steel balls and 410 µL isopropanol, the samples were homogenized using the Geno Grinder 2010, followed by adding 520 µL cyclohexane and vertexing. Phases were then separated by the addition of 570 µL of 0.1 M ammonium acetate and centrifugation for 5 min at 1000 rcf at room temperature. The top 450 µL organic phase was collected, and the aqueous phase was re-extracted with an additional 520 µL of cyclohexane. The combined organic phase was then dried under vacuum and reconstituted in 50 µL toluene followed by 50 µL methanol.

### 2.4 Base hydrolysis of esterified oxylipins

To hydrolyze the complex lipids, the 100 µL of total lipid extract was spiked with 5 µL of 250 nM deuterated Oxyl-EC surrogates in methanol and incubated with 100 µL 0.5 M sodium methoxide for 50 min at 50 °C. Then 100 µL water was followed by another incubation of 50 min at 50 °C. Samples were neutralized by adding 10 µL of 20% glacial acetic acid and then diluted with 0.5 mL 5% methanol - 0.1% acetic acid, as well as 5 µL antioxidant solution. The extracted samples were applied onto a 10 mg Oasis HLB solid phase extraction plate (Water Corp, Milford Mass) to purify non-esterified oxylipins, endocannabinoids, and fatty acids. Next, columns were washed, and analytes were eluted using 0.25 mL of methanol with 1.0% acetic acid and 1 mL ethyl acetate. After being dried under vacuum, samples were reconstituted in 125 µL of 1-cyclohexyl ureido, 3-dodecanoic acid (CUDA), and 1-phenyl ureido 3-hexanoic acid (PUHA) (Cayman Chemical; Ann Arbor, MI) at 100 nM in 1:1 methanol: acetonitrile. Prior to liquid chromatography-mass spectrometry (LC-MS) analysis, samples were filtered through a 0.2 µm PVDF filter plate.

### 2.5 Non-esterified (free) oxylipin, endocannabinoids, and PUFAs

Frozen tissue samples (20 mg ∼ 50 mg) were pre-weighed on dry ice and spiked with 10 µL of 250 nM deuterated Oxyl-EC surrogates in methanol, 5 µL of 5 µM CUDA/PUHA, as well as 5 µL BHT/EDTA at 0.2 mg/ml. To homogenize the samples, 3 mm stainless steel balls and 230 µL 1:1 methanol: acetonitrile were added into each tube before loading the samples into Geo Grinder 2010 for 2 min at 1500 rcf. Prior to LC-MS analysis, samples were centrifuged and filtered through 0.2 µm PVDF filter plate.

### 2.6 Liquid chromatography-mass spectrometry (LC-MS) analysis

Residues in extracts were separated on a 2.1 mm × 150 mm, 1.7 µm BEH C18 column (Waters, Milford, MA) and detected by electrospray ionization with multi-reaction monitoring on an API 6500 QTRAP (SCIEX, Framingham, MA) and quantified against 7 to 9 point calibration curves of authentic standards using modifications of previously reported methods [42]. Concentrations of esterified and non-esterified (free pool) PUFA, oxylipins, endocannabinoids, and a group of non-steroidal anti-inflammatory drugs (NSAIDs) (i.e., ibuprofen and naproxen) were quantified. All samples were processed with quality control measures including randomization, inclusion of method blanks, and analysis of NIST Standard Reference Material 1950 – Metabolites in human plasma (Sigma-Aldrich, St Louis, MO). LC-MS data were processed using MultiQuant v 3.0.3 software (SCIEX). A complete list of analytical parameters are provided in **Appendix A**.

### 2.7 Statistical analysis

All statistical analyses were performed using JMP Student Edition 19.1.3 (SAS Institute, Cary, NC). Prior to statistical analysis, the distribution of each analyte overall and by diagnostic group was characterized, and outliers were assessed, identified, and removed if warranted. Variables were normalized using the distributional Johnson transformations, mean centered, and scaled to a standard deviation of ±1. Analytes with greater than 25% missing values were excluded from the analysis. For the remaining analytes, multivariate normal imputation was used to impute missing values to facilitate statistical analysis. Differences associated with AD diagnosis and sex were evaluated using two-way analysis of covariance (ANCOVA), including AD diagnosis, sex, and the AD-by-sex interaction as fixed factors and analytical batch, age, BMI, and PMI as covariates. ANCOVA models were fitted individually for each analyte and tissue type. For metabolites with a significant AD-by-sex interaction, group-specific metabolite concentrations were examined within each sex to characterize sex-specific effects. Correlations between analytes and BMI or age were assessed using Spearman’s rank-order correlation. The Benjamini– Hochberg procedure was used to control the false discovery rate (FDR) for multiple hypothesis testing; statistical significance was defined at an FDR threshold of 0.20.

For pathway visualization, metabolites showing a significant main effect or interaction were mapped onto corresponding fatty acid and lipid mediator pathways. Mean metabolite concentrations were calculated for each comparison group using the original, untransformed concentration data. For AD-versus-control comparisons, fold changes were calculated as the ratio of group means of the AD relative to the cognitively normal control group; for sex comparisons, fold changes were calculated as the ratio of group means of males relative to females. The magnitude of the difference was represented by node size, whereas the direction of the difference was indicated by node color. Metabolites were further distinguished according to esterified or free lipid fractions. The pathway diagrams were used to visualize observed metabolic differences and did not constitute a separate pathway-enrichment or network statistical analysis.

## 3. Results

### 3.1 Characteristics of the study cohort with paired brain and liver tissues

The basic characteristics of the 79 Caucasian donors are summarized in Table 1. A total of 16 controls and 63 AD cases were included. The mean age was 81.5 and 85.9 years old in cognitively normal (control) and AD groups, respectively. The disparity in sample size between the two groups underscores the intricacies and complexities in the selection of samples from aged donors that were characterized as cognitively normal. Paired prefrontal cortex and liver tissues were sought from each donor in an effort to minimize potential confounding factors from subject differences, however, limitation exists. **Figure 2** provides the age distributions of donors by sex, diagnostic group, and ApoE genotype. Paired brain and liver tissues were obtained from 73 donors while only liver tissues were available from 6 donors. The study cohort was restricted to individuals with homozygous ApoE3/3 or ApoE4/4 genotypes to reduce genetic heterogeneity. Because of the limited and imbalanced genotype subgroup sizes, particularly among ApoE4/4 female donors, ApoE genotype was treated as a descriptive cohort characteristic rather than a primary analytic variable.

**Figure 2.**
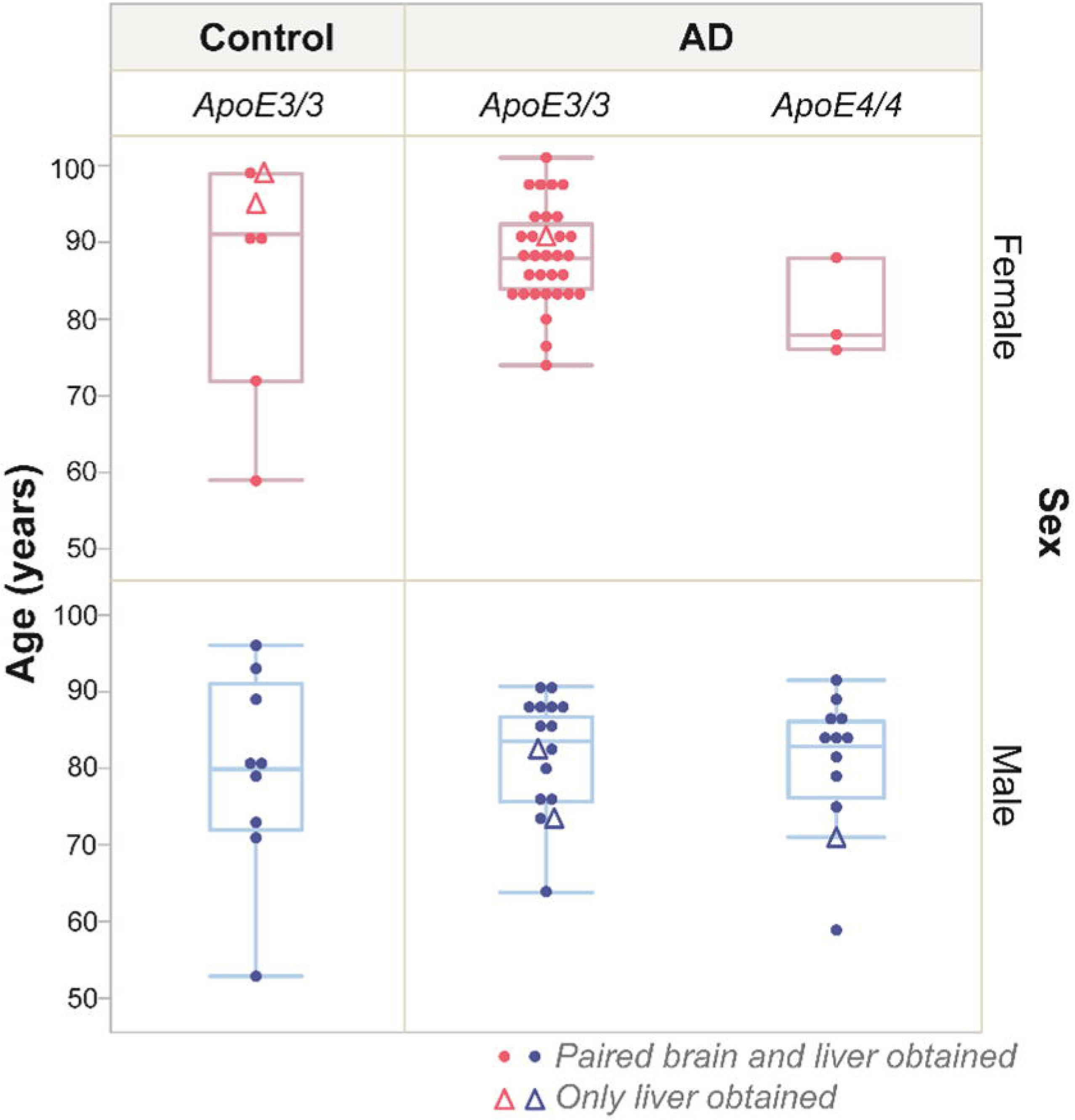
Age distribution of the donors by diagnosis, sex and ApoE genotype. Sex was distinguished by color (pink=female, blue=male). Note that the solid circle dots indicate donors with paired brain and liver tissues (N=73), whereas the triangles indicate donors with only liver tissues available (N=6).

### 3.2 AD is associated with distinct lipid mediator alterations in both brain and liver

The lipid mediator concentrations between controls and AD tissues were compared in 70 oxylipins (of 85 measured in our targeted LC-MS assay), 5 PUFAs (5 measured), 13 endocannabinoids (22 measured), and 2 NSAIDs (4 measured). Lipid mediator concentrations were evaluated using the two-way ANCOVA outlined above. **Table S1** provides the resulting test p-values for the main effects (AD diagnosis, sex), the AD-by-sex interaction, and all adjusting covariates (analytical batch, age, BMI, and PMI). While no AD main effects reached significance after FDR adjustment (FRD < 0.2), metabolites with nominal p-values < 0.05 were leveraged to visualize the overall pattern and scale of AD-associated alternations. Fold changes in the oxylipins, endocannabinoids, and PUFAs were projected onto their corresponding metabolic pathways (**Figure 3**).

**Figure 3.**
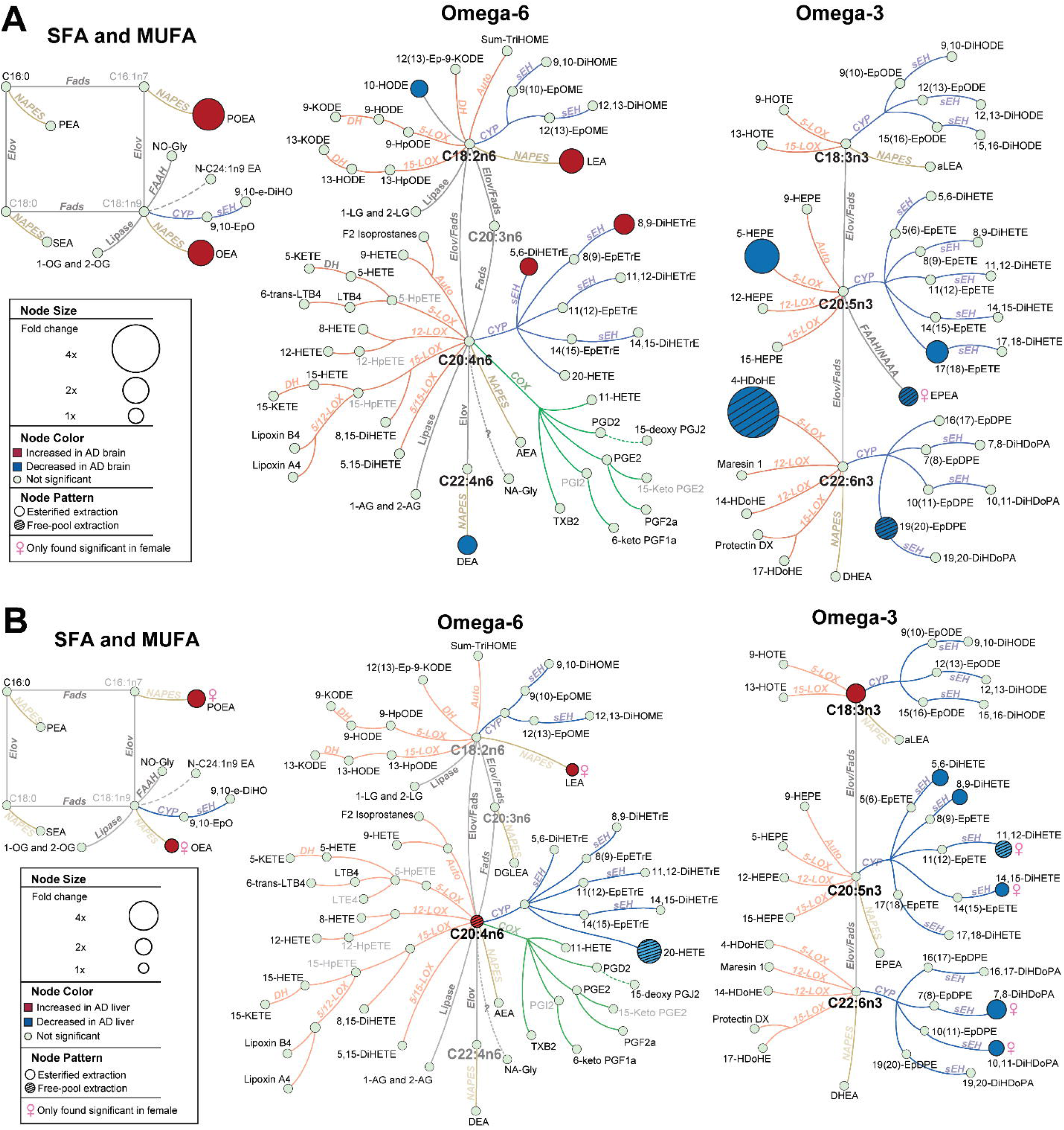
Oxylipin alterations between control (cognitively normal) and AD diagnostic groups observed in brain **(A)** and liver **(B)** tissue. Fold changes reflect the relative magnitude of concentration differences (AD vs. control) across fatty acids, oxylipin, and endocannabinoids metabolic pathways. Only metabolites with a nominal AD main effect (p < 0.05) are displayed, as no alteration researched statistical significance after Benjamini–Hochberg false discovery rate adjustment (FDR < 0.20). The network presents each metabolic pathway involving saturates and monounsaturates (SFA and MUFA), and omega-3 and omega-6 fatty acids with their corresponding oxylipins and endocannabinoids. Detected (black font) and undetected/untargeted (gray font) metabolites were listed to facilitate the visualization and indicate the coverage in our targeted LC-MS assay. Although, to simplify the network mapping, not all metabolites were displayed. Oxylipin metabolizing enzymes were color coded into classifications. Specifically, orange shows lipoxygenase (LOX) and autoxidation pathway; blue represents cytochrome p450 (CYP)/soluble epoxide hydrolase (sEH); green shows cyclooxygenase (COX); yellow means N-acylphosphatidylethanolamide-phospholipase D. Node size represents the absolute fold change, and color denotes the direction of change (red: higher in AD; blue: lower in AD). Node patterns distinguish oxylipin fractions, where a solid fill indicates esterified extraction, and a lined filling pattern indicates free-pool extraction. The female symbol (♀) identifies metabolites with a significant AD-by-sex interaction, where the AD-associated alternation was observed in females. Key enzymes abbreviation: Fads, fatty acid desaturase; Elov, fatty acid elongase; DH, dehydrogenase. SFA and MUFA were not measured in the assay but were indicated only to visualize precursors for measured oxylipins and endocannabinoids. The resulting ANCOVA p-values, including AD-by-sex interaction terms, are provided in **Table S1**.

As shown in **Figure 3A**, AD-associated alternations were observed across multiple fatty acid-derived lipid mediator pathways in brain tissue. EPA- and DHA-derived monoalcohols, including the 5-LOX-derived metabolites 5-HEPE and 4-HDoHE, were approximately 3.5-fold lower in AD than in controls. CYP-derived metabolites from EPA and DHA, including 17(18)-EpETE and 19(20)-EpDPE, were also lower in AD brain. In contrast, higher levels of acylethanolamides derived from LA (LEA), palmitic acid (POEA), and oleic acid (OEA) were observed in AD, whereas the docosatetraenoic acid-derived ethanolamide DEA was lower. Non-esterified EPEA exhibited a significant AD-by-sex interaction, characterized by lower concentrations in AD females than in cognitively normal females.

AD-associated alternations in liver tissue were similarly projected onto the corresponding metabolic pathways (**Figure 3B**). While several acylethanolamide metabolic alternations were conserved between tissues, the liver exhibited a distinctively broader profile of metabolic shifts compared to the brain. Significant AD-by-sex interactions were observed for POEA, OEA, and LEA, revealing higher levels in AD females relative to cognitively normal females. PUFA concentrations, including AA and αLA, were higher in AD liver. The largest alteration was observed for 20-HETE, a CYP-derived metabolite of AA, which was approximately 3.2-fold lower in AD than in controls. sEH-derived metabolites from EPA and DHA were also lower in AD liver. Several dihydroxy fatty acids, including 11,12-DiHETE, 14,15-DiHETE, 7,8-DiHDoPA, and 10,11-DiHDoPA, showed significant AD-by-sex interactions, revealing lower levels in AD females than in cognitively normal females.

### 3.3 Sex is associated with distinct lipid mediator profiles in brain and liver

Given previously reported sex differences in lipid metabolism [43], we further assessed sex as a main effect on lipid mediator profiles, controlling for AD diagnostic status and other covariates. As shown in **Figure 4**, metabolites showing significant sex-associated differences (FDR < 0.20) were mapped onto their corresponding metabolic pathways, with effect magnitudes expressed as male-to-female abundance ratios. The resulting ANCOVA p-values for the main effect of sex and other covariates, are provided in **Table S2**. Sex-associated alternations were more pronounced in liver than in brain. In the liver (**Figure 4B**), several LOX-derived metabolites from AA and LA were higher in males than females. In contrast, free EPA-derived epoxides and esterified diols derived from EPA and αLA were lower in males than in females. Fewer sex-associated alterations were observed in brain tissue (**Figure 4A**); among these, 8,15-DiHETE, 11-HETE, and 15,16-DiHODE were elevated in males compared to females.

**Figure 4.**
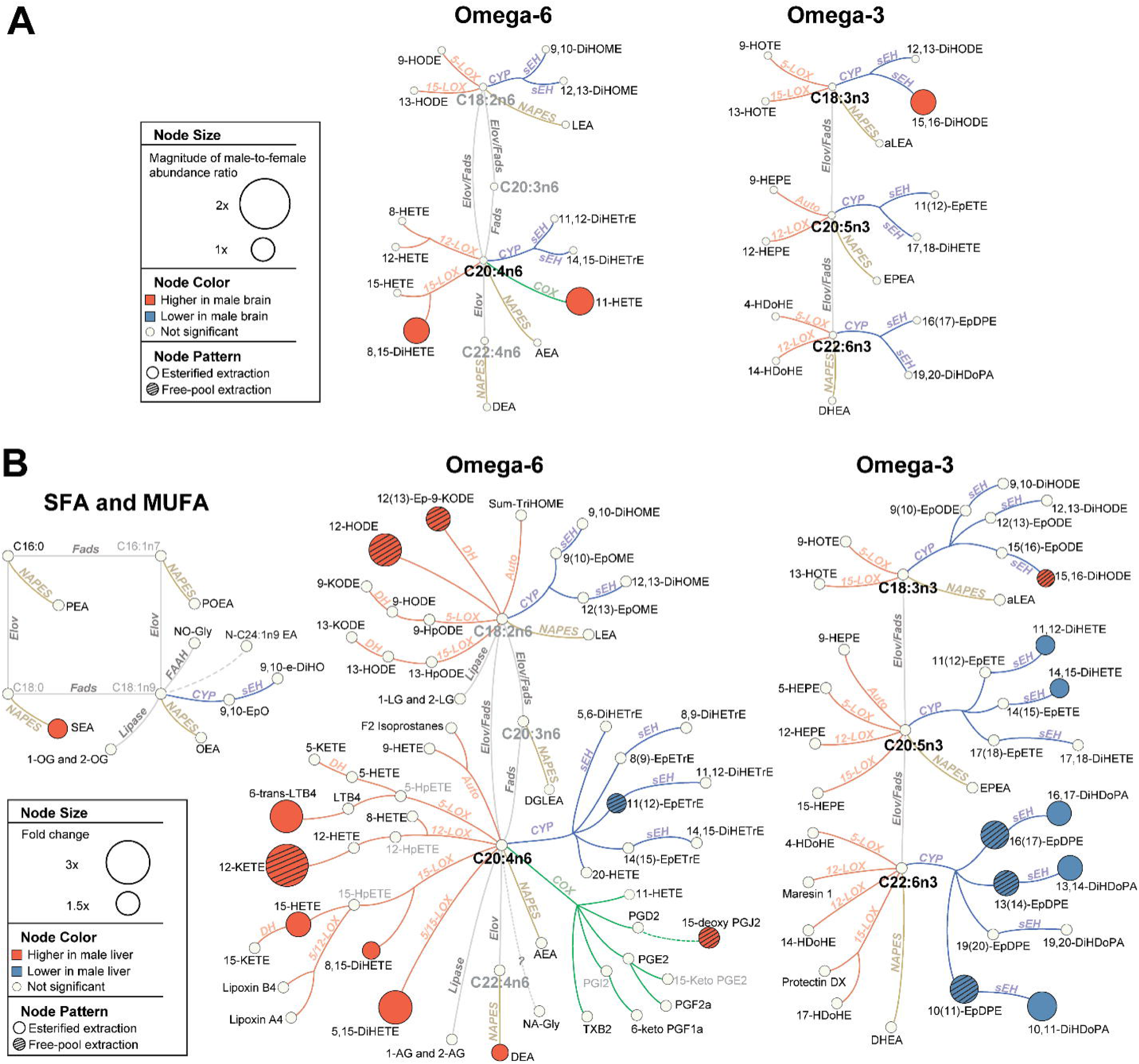
Sex-associated alternations in lipid mediator profiles in brain **(A)** and liver **(B)**. Fold changes represent male-to-female abundance ratios. Displayed metabolites exhibited a significant main effect of sex (FDR < 0.2) in ANCOVA models, adjusted for AD diagnosis, analytical batch, and age. SFA, MUFA, omega-3, and omega-6 fatty acids are presented with their corresponding oxylipins, endocannabinoids, and metabolic pathways. The brain network **(A)** was simplified for illustrative clarity because of fewer significant sex-associated alternations. Node size reflects absolute fold change, and node color denotes the direction of change (orange: higher in males; blue: lower in males). Node patterns designates the targeted lipid fractions (solid fill: esterified fraction; line pattern: free fraction). Effect-test p-values for the main effect of sex and corresponding model covariates are provided in **Table S2** provides the resulting ANCOVA p-values for the sex main effects and accompanying covariates.

## 4. Discussion

In this study, we performed a comprehensive analysis of both esterified and free oxylipins and endocannabinoids in paired postmortem brain and liver tissues from cognitively normal and AD donors. The short PMI (median of 3.08 hours) and availability of paired tissues provided a unique opportunity to examine central and peripheral lipid metabolism within the same individuals. Our findings demonstrate distinct AD-associated alterations in lipid mediator metabolism in both brain and liver, with a more extensive pattern of divergence observed in liver. Particularly, alterations in CYP/sEH-related metabolites and fatty acid ethanolamides were prominent across tissues, and several metabolites exhibited significant AD-by-sex interactions. Together, these findings support the concept that AD-associated lipid dysregulation extends beyond the brain and involves peripheral metabolic pathways, underscoring a significant role for the liver.

Previous studies examining plasma and CSF have implicated oxylipins, endocannabinoids, and related lipid pathways in AD-associated inflammatory processes [14]. In plasma, the sEH EPA metabolite 17,18-DiHETE was elevated approximately 3-fold in AD, whereas acylethanolamides derived from several fatty acids were significantly reduced. In CSF, fewer lipid metabolites were detected, with the most pronounced alterations occurred within the LA CYP/sEH pathway, where both epoxides and diols were higher in the AD group compared to the controls [14]. Large-scale studies from the Alzheimer’s Disease Metabolomics Consortium have further linked CYP/sEH and fatty acid ethanolamide metabolism to AD pathology, cognitive decline, and resilience to tau and Aβ pathology [8]. Integration of metabolomic and proteomic datasets has also linked these pathways to vascular inflammation and energy metabolism within the central nervous system [44, 45]. Our tissue-based findings extend this literature, demonstrating that AD-associated alterations in these lipid mediator pathways are also detectable directly within human brain and liver tissues and are not restricted to circulating or cerebrospinal fluid compartments.

An important feature of the present study is the simultaneous characterization of esterified and non-esterified lipid mediator pools. Previous investigations have primarily focused on free oxylipins in biofluids, even though esterified lipid mediators constitute a substantial reservoir within complex lipids. In contrast to prior plasma observations showing lower free fatty acid ethanolamides in AD [14], we observed higher levels of several esterified ethanolamides in both AD brain and liver (**Figure 3**). These differences suggest that AD-associated lipid dysregulation may involve not only changes in bioactive free mediators but also alterations in their storage, incorporation, and mobilization within complex lipid pools. Because esterified oxylipins undergo continuous release and re-esterification, changes in this compartment may reflect altered availability of precursors for subsequent lipid signaling. However, the present cross-sectional tissue measurements cannot establish the directionality or kinetics of exchange between esterified and free lipid fractions. Thus, rather than indicating that esterified alterations necessarily precede changes in the free pool, our findings demonstrate that these two fractions provide complementary information about AD-associated lipid homeostasis.

Differences between tissue and circulating lipid pools were particularly apparent for CYP/sEH-related metabolites. Decreased esterified diols were observed in AD liver, whereas previous plasma studies reported increased circulating diols associated with the sEH pathway [14, 46]. These apparently discordant patterns may reflect complex differences in lipid compartmentalization, tissue-specific metabolism, and exchange between esterified and free pools. Esterified oxylipins function as dynamically remodeled reservoirs that can undergo hydrolysis and re-esterification, and therefore tissue concentrations do not necessarily parallel circulating abundance [23, 47–49]. While the absence of matched plasma samples from these donors prevents a direct assessment of whether tissue and circulating pools are inversely or coordinately regulated, these contrasting patterns emphasize the importance of examining both lipid fraction and tissue compartment when interpreting oxylipin metabolism in AD.

The extensive metabolic alterations observed in liver are particularly notable given its central role in systemic lipid metabolism. Increased ethanolamides together with alterations in CYP/sEH-related metabolites suggest substantial remodeling of hepatic lipid mediator homeostasis in AD. Our findings identify hepatic metabolism as a potentially important peripheral component of AD-associated lipid dysregulation, and it would be important to test whether some of the liver lipid changes may drive brain lipid changes,. The liver regulates fatty acid metabolism, lipoprotein production, and circulating lipid mediator availability and therefore provides a biologically plausible interface between systemic metabolic conditions and the brain [50]. This possibility of inter-organ communication is supported by experimental models; for instance, the modulation of hepatic sEH in mice has been shown to alter systemic inflammatory lipid profiles and cerebral Aβ metabolism and influence AD-related pathology [51]. Our human tissue observations therefore complement these mechanistic animal studies and provide further rationale for investigating hepatic lipid mediator metabolism as part of a broader brain–periphery metabolic axis in AD. Direct correlation analyses of matched brain-liver metabolite profiles will be needed to determine whether specific lipid mediator pathways demonstrate coordinated regulation across tissues.

Sex was also associated with distinct lipid mediator profiles, a phenomenon that was particularly pronounced in liver. Importantly, these findings represent the main effect of sex across the cohort, distinct from the AD-by-sex interactions identified for specific metabolites in the AD analyses. Males exhibited higher levels of several LOX-derived metabolites from AA and LA, consistent with previous evidence of sex-related alternations in eicosanoid formation and metabolism [52]. Given LOX-derived lipid mediators have also been implicated in cardiovascular and vascular inflammatory processes [53], these findings may be relevant to sex-dependent differences in systemic inflammatory and vascular biology. In contrast, free EPA-derived epoxides were depleted in males. Previous studies have similarly reported sex differences in long-chain PUFA metabolism and status, including higher hepatic and circulating DHA levels in females than in males and sex-specific efficiency in converting αLA to longer-chain n-3 PUFAs [54–56]. Such differences in precursor fatty acid metabolism may contribute to the sex-associated patterns of downstream oxylipins observed here. Since epoxy fatty acids have been associated with anti-inflammatory and pro-resolving signaling, lower concentrations in males may be consistent with sex-specific alternations in inflammatory lipid regulation; however, metabolite concentrations alone do not establish underlying differences in enzymatic activity. Esterified EPA- and αLA-derived diols were also lower in male liver, potentially reflecting sex-dependent differences in lipid incorporation, storage, or remodeling dynamics. Fewer sex-associated alternations were observed in brain; however, 8,15-DiHETE, 11-HETE, and 15,16-DiHODE – metabolites known to be associated with inflammatory lipid pathways - were elevated in males. Previous studies have reported sex differences in AD-related vulnerability and progression, including divergent relationships between AD biomarkers and cognitive decline, hippocampal atrophy, and brain volume loss [57, 58]. As precision medicine approaches continue to develop, biological sex is increasingly recognized as an important source of heterogeneity in AD. Nonetheless, the metabolic mechanisms underlying these differences remain incompletely understood and are likely multifactorial [59–61].

Altogether, these findings support a model in which AD is characterized by both central and peripheral dysregulation of lipid mediator metabolism across brain and liver, with hepatic metabolic alterations potentially contributing to or reflecting central inflammatory lipid remodeling. Analyses examining correlations between brain and liver lipid mediator profiles are currently underway to better understand potential metabolic coordination between the central and peripheral systems. Future studies incorporating longitudinal biofluids together with tissue measurements will also be particularly important to establish how these pools relate and to determine whether tissue esterified lipid profiles have predictive value as indicators of longer-term metabolic remodeling.

Several limitations should be considered when interpreting our findings. First, while the use of paired brain and liver tissues represents a major strength, the relatively small size of the cognitively normal group and the scarcity of well-characterized postmortem specimens limited our statistical power for subgroup analyses, including detailed evaluation of ApoE genotype. Consequently, ApoE genotype was treated primarily as a descriptive cohort characteristic rather than a major analytic focus in the present study. Second, all donors were of Caucasian background, which restricts generalizability of these findings across the broader ethnoracial heterogeneity characteristic of AD. Future studies in larger, more diverse cohorts will be essential to determine whether these metabolic patterns remain consistent across varied demographic and genetic backgrounds. Third, matched plasma or CSF samples were not available for this cohort, precluding a direct comparison between tissue and circulating lipid mediator pools. Multi-tissue studies integrating matched peripheral biofluids with brain and peripheral tissues will be needed to define lipid mediator exchange and coordination across compartments. Finally, the observational and cross-sectional nature of this postmortem study limits our ability to determine causal inferences regarding whether the observed lipid alterations contribute to or manifest as a consequence of AD pathology.

In conclusion, this study identifies distinct AD-associated lipid mediator profiles across paired human brain and liver tissues, highlighting alterations in CYP/sEH-related metabolism, fatty acid ethanolamides, and esterified lipid mediator pools. The more extensive alterations observed in liver support the importance of considering peripheral lipid metabolism alongside central nervous system processes when investigating AD mechanisms. Our findings also demonstrate substantial sex-dependent variation in lipid mediator metabolism, particularly in liver. Together, these observations provide a framework for investigating coordinated central and peripheral lipid dysregulation in AD and support future studies that integrate brain, liver, and circulating lipid mediator profiles to determine their mechanistic roles and biomarker potential.

## Data Availability

Additional information is available in Supporting Information or upon reasonable request.

## Acknowledgement

This work was supported by the National Institute on Aging (RF1AG071665 and P30 AG072972), Eunice Kennedy Shriver National Institute of Child Health and Human Development (P50HD103526). Additional support was provided by USDA Project 2032-10700-003-000-D. The USDA is an equal opportunity provider and employer. We gratefully acknowledge Dr. Bruce D. Hammock for valuable insights, initial study conceptualization, and thoughtful guidance throughout its development. We sincerely thank the donors and their families for their essential contributions. We thank Dr. Isabel F. Snodgrass for her guidance and assistance during instrument operation and data acquisition. We also acknowledge the Banner Sun Health Research Institute Brain and Body Donation Program (Sun City, Arizona) for providing human biological materials. The Brain and Body Donation Program has been supported by the National Institute of Neurological Disorders and Stroke (U24 NS072026 National Brain and Tissue Resource for Parkinson’s Disease and Related Disorders), the National Institute on Aging (P30 AG019610 and P30AG072980, Arizona Alzheimer’s Disease Center), the Arizona Department of Health Services (contract 211002, Arizona Alzheimer’s Research Center), the Arizona Biomedical Research Commission (contracts 4001, 0011, 05-901 and 1001 to the Arizona Parkinson’s Disease Consortium) and the Michael J. Fox Foundation for Parkinson’s Research.

## Consent Statement

All study subjects provided informed consent prior to participation in the study.

