## Supplemental Tables for "Distinct lipid mediator profiles in brain and liver tissues in Alzheimer’s disease"

**Table S1.** ANCOVA p-values of significant metabolites associated with main effects (AD and Sex) and AD-by-Sex interactions (Sex*AD), while controlling for covariates (PMI, batch, age, and BMI), with the exclusion of ApoE4/4 samples.

| **Metabolites** | **Tissue Type** | **AD** | **Batch** | **Age** | **Sex** | **Sex*AD** | **BMI** | **PMI** |
| --- | --- | --- | --- | --- | --- | --- | --- | --- |
| Free-11,12/14,15-DiHETrE | Liver | 0.0091 | 0.0523 | 0.2156 | 0.7298 | 0.7439 | 0.8104 | 0.2198 |
| Free-14,15/11,12-DiHETrE | Liver | 0.0103 | 0.0516 | 0.2408 | 0.7106 | 0.7627 | 0.8126 | 0.2239 |
| Bound-N-C20:5n3_EA | Brain | 0.0131 | 0.2197 | 0.1879 | 0.5991 | 0.0724 | 0.3254 | 0.2365 |
| Bound-10-HODE | Brain | 0.0137 | 0.0248 | 0.1287 | 0.9453 | 0.5047 | 0.1021 | 0.0349 |
| Free-20-HETE | Liver | 0.0182 | 0.4951 | 0.9492 | 0.3441 | 0.4536 | 0.5313 | 0.8382 |
| Free-Sum(DiHOME)/Sum(EpOME) | Liver | 0.021 | 0.1752 | 0.9307 | 0.5597 | 0.8002 | 0.3057 | 0.6717 |
| Free-N-C22:4n6_EA | Brain | 0.0229 | 0.0128 | 0.2408 | 0.2784 | 0.2921 | 0.4802 | 0.2339 |
| Bound-17-KDoHE | Brain | 0.026 | 0.0012 | 0.0285 | 0.4387 | 0.9111 | 0.1425 | 0.9389 |
| Bound-5,6-DiHETE | Liver | 0.027 | 0.0404 | 0.0293 | 0.5297 | 0.0041 | 0.5301 | 0.7885 |
| Free-11,12-DiHETE | Liver | 0.0284 | 0.3621 | 0.1051 | 0.9747 | 0.5844 | 0.9639 | 0.049 |
| Bound-17(18)-EpETE | Brain | 0.0289 | <.0001 | 0.3781 | 0.9671 | 0.2038 | 0.2908 | 0.4025 |
| Bound-8,9-DiHETE | Liver | 0.0343 | 0.5383 | 0.5941 | 0.176 | 0.0002 | 0.6138 | 0.7165 |
| Bound-8,9-DiHETrE | Brain | 0.0366 | 0.0341 | 0.6428 | 0.0586 | 0.0444 | 0.8113 | 0.6229 |
| Bound-5,6-DiHETrE | Brain | 0.0378 | 0.0018 | 0.4037 | 0.054 | 0.0744 | 0.1906 | 0.979 |
| Bound-C18:3n3 | Liver | 0.041 | <.0001 | 0.1274 | 0.6497 | 0.405 | 0.202 | 0.9415 |
| Bound-EPA + DHA diols | Brain | 0.0429 | 0.02 | 0.631 | 0.4161 | 0.0553 | 0.4766 | 0.1022 |
| Bound-18-HEPE | Brain | 0.0431 | 0.3432 | 0.8775 | 0.4679 | 0.4159 | 0.8376 | 0.2814 |
| Free-C20:4n6 | Liver | 0.0458 | <.0001 | 0.1495 | 0.8376 | 0.7687 | 0.8056 | 0.8495 |
| Free-19(20)-EpDoPE | Brain | 0.0475 | 0.0334 | 0.622 | 0.7215 | 0.3916 | 0.2931 | 0.0958 |
| Bound-4-HDoHE | Brain | 0.0493 | 0.0177 | 0.7666 | 0.1031 | 0.7339 | 0.7439 | 0.9314 |
| Bound-5-HEPE | Brain | 0.0498 | 0.7472 | 0.4322 | 0.1533 | 0.1446 | 0.8235 | 0.8601 |
| Bound-14,15/17,18-DiHETE | Brain | 0.0526 | 0.0702 | 0.8066 | 0.6457 | 0.8812 | 0.9124 | 0.0382 |

**Table S2.** ANCOVA p-values of metabolites showing significant sex differences in lipid mediators, adjusted for AD diagnosis, analytical batch, and age.

| **Metabolites** | **Tissue Type** | **P-value** | **P-value** | **P-value** | **P-value** |
| --- | --- | --- | --- | --- | --- |
|  |  | **AD vs Control** | **Batch** | **Age** | **Sex** |
| **Free-12(13)-Ep-9-KODE** | Liver | 0.4264 | 0.8614 | 0.0897 | 0.0025 |
| **Free-13(14)-EpDoPE** | Liver | 0.7158 | 0.2449 | 0.15 | 0.004 |
| **Free-15-deoxy_PGJ2** | Liver | 0.6127 | 0.8202 | 0.6365 | 0.0101 |
| **Bound-5,15-DiHETE** | Liver | 0.3314 | 0.9282 | 0.3483 | 0.0147 |
| **Free-11(12)-EpETrE** | Liver | 0.9287 | 0.7909 | 0.3219 | 0.0153 |
| **Bound-6-trans-LTB4** | Liver | 0.9768 | 0.2077 | 0.6179 | 0.0154 |
| **Bound-N-C20:4n6_Ser** | Liver | 0.0976 | 0.0633 | 0.0159 | 0.0164 |
| **Bound-8,15-DiHETE** | Brain | 0.6983 | 0.0004 | 0.2444 | 0.0166 |
| **Free-15,16-DiHODE** | Liver | 0.2862 | 0.2488 | 0.4284 | 0.0172 |
| **Bound-15-HETE** | Liver | 0.3381 | 0.9505 | 0.0388 | 0.0177 |
| **Free-Acetaminophen** | Liver | 0.1408 | 0.5562 | 0.1813 | 0.0195 |
| **Bound-10,11-DiHDoPE** | Liver | 0.2373 | 0.0003 | 0.0388 | 0.0239 |
| **Bound-11,12-DiHETE** | Liver | 0.2064 | 0.8692 | 0.3375 | 0.0242 |
| **Bound-13,14-DiHDoPE** | Liver | 0.4385 | 0.6624 | 0.1913 | 0.0259 |
| **Free-10(11)-EpDoPE** | Liver | 0.8577 | 0.2739 | 0.382 | 0.0325 |
| **Bound-N-C18:0_EA** | Liver | 0.7748 | 0.0713 | 0.7734 | 0.0335 |
| **Bound-N-C16:0_Gly** | Liver | 0.9561 | 0.0142 | 0.7182 | 0.0395 |
| **Bound-11-HETE** | Brain | 0.4463 | 0.0007 | 0.9071 | 0.0417 |
| **Bound-16,17-DiHDoPE** | Liver | 0.6109 | 0.283 | 0.2882 | 0.0422 |
| **Bound-N-C22:4n6_EA** | Liver | 0.4182 | 0.0409 | 0.4251 | 0.0432 |
| **Bound-15,16-DiHODE** | Brain | 0.8895 | <.0001 | 0.5163 | 0.0453 |
| **Free-16(17)-EpDoPE** | Liver | 0.9899 | 0.5405 | 0.5038 | 0.046 |
| **Free-12-HODE** | Liver | 0.5816 | 0.8881 | 0.6454 | 0.0464 |
| **Bound-14,15-DiHETE** | Liver | 0.5097 | 0.8628 | 0.3435 | 0.0479 |
| **Free-12-KETE** | Liver | 0.8739 | 0.1639 | 0.216 | 0.0495 |
